# Growing into Perception: Pubertal Stages and the Accuracy of Body Size Estimation

**DOI:** 10.1101/2025.11.13.25340184

**Authors:** Maurício José de Siqueira Cagno, Tatiana Wscieklica, Eliane Florencio Gama

## Abstract

Puberty is a significant period of physical, psychological, and social transformation. During this phase, rapid changes in body size, shape, and composition occur, largely regulated by endocrine factors, and are accompanied by shifts in body perception. Body image, a multidimensional construct influenced by biological, psychological, and socio-cultural factors, plays a crucial role in adolescent health and development. The perception of body size, a key aspect of body image, has been found to change during adolescence, influenced by factors such as weight gain, personal relationships, and media exposure. While much of the existing literature on adolescent body image focuses on cognitive, emotional, and attitudinal perceptions, there is limited research on the dimensional aspect of body image, particularly in relation to puberty-related somatic transformations. This study aimed to assess the effect of puberty-related somatic changes on adolescents’ body size estimation across different stages of sexual maturation. We conducted a cross-sectional study with 100 pubescent participants, aged 9 to 15 years for males and 9 to 13 years for females, who were assessed using the Image Marking Procedure (IMP) to estimate body size perception. Our data suggest that adolescents at an early stage of sexual maturation, have an overestimated perception of their body size and this distortion seems to get closer to adequate as sexual maturity develops. The general perception of adolescents’ body size is overestimated in girls and adequate in boys. This research underscores the evolving nature of body size perception during puberty and its potential implications for adolescent body image.

**Highlights:**

- Puberty changes affect adolescents’ body size perception across maturation stages.
- Early pubertal stages are associated with body size overestimation, which tends towards accuracy with advancing sexual maturation
- Overall body size perception is overestimated in girls and adequate in boys.

## 1.1 Introduction

Puberty is a period of significant physical maturation and accompanying social and behavioral changes, typically occurring between 8 and 24 years of age, with variations influenced by sex, ethnicity, health status, genetics, nutrition, and activity level (Pinyerd & Zipf, 2005; Sawyer et al., 2018). The period from the onset of puberty to the completion of physical maturation is often referred to as adolescence (Rosenfeld, 1982) a transitional phase from childhood to adulthood characterized by ongoing physical, psychological, and social development (Sawyer et al., 2018).

During puberty, rapid and substantial changes in body size, shape, and composition occur, largely regulated by endocrine factors (Loomba-Albrecht & Styne, 2009; Rogol et al., 2002). Interestingly, the growth spurt can follow a cyclical pattern, with limb size and longitudinal height developing at varying velocities (Butler et al., 1990; Hermanussen et al., 1998). Given these rapid physical transformations, adolescents’ perception of their body dimensions is also likely to evolve throughout early and middle adolescence (Tadai et al., 1994). A longitudinal study among Italian adolescents, for instance, showed that girls tended to overestimate their body size (Toselli et al., 2021).

The perception of body size, which refers to an individual’s awareness and judgment of their physical dimensions, can be understood as the dimensional aspect of body image. As a dynamic and multidimensional construct, body image is shaped by a complex interplay of perceptions, emotions, behaviors, beliefs, external influences, and past experiences (Bonev & Matanova, 2021; Cash, 2012; Tylka & Wood-Barcalow, 2015). The development of body image during adolescence is critically shaped by the various biological (e.g., weight gain), psychological (e.g., personal relationships), and socio-cultural factors (e.g., media) that accompany this period (Hartman-Munick et al., 2020), playing an important role in adolescent health and development. While extensive research has explored the attitudinal, cognitive, and emotional facets of body image in adolescents (Markey, 2010).the dimensional aspect, particularly in relation to puberty-driven somatic changes, remains understudied. This gap in the literature motivates the present study, which aims to assess the effect of puberty-related somatic transformations at different stages of sexual maturation on body size estimation.

## 2. Methods

### 2.1 Participants

This study involved 100 pubescent children whose legal representatives or guardians authorized on their behalf by signing the Informed Consent Form number and was approved by the ethical committee from Universidade São Judas Tadeu, number 089/11. We assessed 100 pubescent, 50 males aged between 9 and 15 years old and 50 females aged between 9 to 13 years old, with subgroups of ten participants from each age group. All the individuals invited to take part in this research were enrolled in elementary school of a regular school in São Paulo, located in the urban but peripheral region of the East Zone. During each school year, the subjects took part in physical education classes, twice a week, for a duration of 45 minutes each. We also investigated the accumulated months of Previous Motor Activities (extracurricular sports or recreational motor activities practiced from childhood until the time of data collection) and the weekly hours of Global Motor Inactivity (time per week spent sitting in front of the television, computer sitting in front of the television, computer, video game and similar without performing activities) in order to analyze the possible influences of these variables on the results in self-filled questionnaire.

### 2.2 Sexual maturation

Participants had to be in the sexual maturation stage from I to IV, according to the self-administered instrument (Morris & Udry, 1980). The use of sexual maturation was a necessary procedure to consider since, by taking only age in account, there could be a bias in the samples. There could be an early or late pubescents within a more advanced or delayed chronological age.

### 2.3 Body image dimension

The perception of body size can be investigated using analogue scales, image marking and optical distortion methods (Farrell et al., 2005). The Image Marking Procedure (IMP) was chosen as it is an extremely simple and inexpensive test. The researcher placed stickers on the participants clothes so that they don’t move out of place and the researcher would always touch the same place, in seven body landmarks: top of the head, left acromio-clavicular joint, right acromio-clavicular joint, narrowest waist width from both sides, left trochanter and right trochanter (Askevold, 1975). This test has been used in prior literature (Picado et al., 2019; Pivotto et al., 2022; Souza et al., 2023; Thurm et al., 2013).

Then participants were asked to imagine themselves standing before a mirror and looking at themselves, and then close their eyes. The examiner, behind the participant, touched the body landmark with his fingers and then the participant was instructed to touch the wall in front of himself, while the researcher marked the wall with a sticker as if he was projecting the location of the body landmarks in the mirror. For the top of the head, participants were instructed to hold their breath, in positive apnea. This procedure was done three times with the participant using a blindfold. Then, participants would approach the wall, maintaining the anterior foot alignment. and the real dimensions were marked using a ruler, the labels and measuring tape on the wall were photographed with a digital camera for further analysis (Fonseca et al., 2014; Thurm et al., 2013)

#### Calculations

The distances of the points identified by the participant and the real dimensions of height and width in the horizontal plane were assessed. Data were organized in tables and the overall Body Perception Index (oBPI) was calculated. First, the BPI of each region was analyzed. From the three attempts of perceived measures for each region Perceived size was calculated as the average response divided by actual measurement, multiplied by 100. Overall BPI (oBPI) [(head BPI + shoulder BPI + waist BPI + hip BPI) / 4] and head BPI [head BPI = (mean head BPI / real head BPI)*100 were calculated ( Askevold, 1975 ; Fonseca, 2009; Segheto, et al., 2011 ; Thurm, et al., 2013). The classification of oBPI followed Segheto, et al. (2011), who suggested a classification standard of general body awareness of individuals. This classification was based on the percentile measurements of adults, where < 99,4% is considered as underestimated, 99,4% - 112,3% is considered adequate and > 112,3% is considered overestimated (Segheto et al., 2011). It is important to note that the classification was made based on adults’ body perception index, as there were no articles with a standardization in adolescents population.

### 2.4 Data analysis

For the statistical analysis, a one-way ANOVA was conducted to compare the effects of sexual maturation on the studied variables. Prior to performing ANOVA, the assumptions of normality and homogeneity of variances were assessed using the Shapiro-Wilk test and Levene’s test, respectively. When these assumptions were violated (p < 0.05), the raw data were transformed into z-scores to standardize the distributions and mitigate potential biases due to differences in scale and variability. The z-score transformation maintains the relative differences between data points while allowing comparisons on a standardized scale with a mean of zero and a standard deviation of one. Despite the transformation, if the data still did not meet ANOVA assumptions, this limitation was acknowledged in the interpretation of results. Frequency and percentage tables for discrete variables and the mean were also described. Partial eta squared (η²p) was used as a measure of effect size. All analyses were performed using JAMOVI software, with a significance level set at α = 0.05.

## 3. Results

We first analysed the participants oBPI and classified them as underestimated, adequate and overestimated. The data reveals important nuances in the distribution. The underestimated group consisted of 12 individuals (n = 7 male and n = 5 female, M = 96.3, SD = 2.61), the group with adequate perception consisted of 48 individuals (n = 29 male and n = 19 female, M = 106, SD = 3.6), and the overestimated group consisted of 40 participants (n = 16 male and n = 24 female, M = 124, SD = 9.98). We found out that the general perception of adolescents’ body size is overestimated in girls (mean = 114, SD = ± 14.3) and adequate in boys (mean = 110, SD = ± 9.43) in our sample.

A ANOVA examines the effects of classification (underestimated, adequate and overestimated), sex (female, male), sexual maturation (stages I, II, III and IV) and their interaction on oBPI. Assumption checks revealed a violation of homogeneity of variances [Levene’s test: F(2,97) = 11.5, p < 0.001] and normality [Shapiro-Wilk: W = 0.861, p < 0.001] for classification, [Levene’s test: F(1,98) = 7.89, p = 0.006] and normality [Shapiro-Wilk: W = 0.955, p = 0.002] sex, [Levene’s test: F(3,96) = 3.39, p = 0.021] [Shapiro-Wilk: W = 0.959, p = 0.003] and for sexual maturation, transformed the raw oBPI data in z-score and then performed the ANOVA (Zscore_oBPI).

Classification shows a highly significant effect (F(2, 97) = 108, p < .001, η² = 0.691), indicating substantial differences in “Zscore_oBPI” across the classification groups. Post hoc Tukey tests revealed significant differences between all classification groups. The overestimated group had significantly higher oBPI scores compared to both the adequately estimated (mean difference = 17.6, SE = 1.47, p < 0.001) and underestimated groups (mean difference = 27.7, SE = 2.26, p < 0.001). Additionally, the adequately estimated group presented significantly higher scores than the underestimated group (mean difference = 10.0, SE = 2.21, p < 0.001). “Sex” does not exhibit a significant effect (F(1,98) = 2.82, p = 0.096, η² = 0.028), suggesting no significant difference in “Zscore_oBPI” between genders. The analysis reveals that sexual Maturation does not have a statistically significant effect on oBPI (F(3, 96) = 1.573, p = 0.194, η²p = 0.048). This indicates that there are no significant differences in oBPI across the different levels of sexual maturation.

The distribution of the analyzed variable across the four stages exhibited distinct characteristics in terms of central tendency, dispersion, and normality (Table 1). Stage I, composed of a small sample (N=7) exclusively of females, presented a mean of 120 (95% CI: 99.3–140) and a median of 109, suggesting a potentially asymmetrical distribution. The relatively high standard deviation (22.2) indicated greater variability within this group. The Shapiro-Wilk test was non-significant (p = 0.105), suggesting that the data may follow a normal distribution. Stage II, with a moderate sample size (N=17), showed a mean of 115 (95% CI: 109–120), a median of 114, and a lower standard deviation (10.9), indicating a more homogeneous distribution. The Shapiro-Wilk test was also non-significant (p = 0.968), reinforcing the assumption of normality. Similarly, Stage III (N=19) presented a mean of 113 (95% CI: 107–118), a median of 115, and a standard deviation of 11.5, with a non-significant Shapiro-Wilk test (p = 0.587), suggesting normal distribution. In contrast, Stage IV, the largest sample (N=57), had a mean of 110 (95% CI: 107–113), a median of 108, and a standard deviation of 11.0. Unlike the previous stages, the Shapiro-Wilk test was significant (p < 0.001), indicating a deviation from normality in this group. In summary, sexual maturation does not appear to have a significant effect on oBPI.

**Table 1.** Shows the number of participants in each stage of sexual maturation, mean oBPI, confidence interval and standard deviation of the different stages of sexual maturation.

|  | Sexual<br>Maturatio<br>n | N | Mean | SD | Min | Max |
| --- | --- | --- | --- | --- | --- | --- |
| oBPI | I | 7 | 120 | 22.2 | 101.2 | 162 |
|  | II | 17 | 115 | 10.9 | 95.9 | 137 |
|  | III | 19 | 113 | 11.5 | 91.3 | 130 |
|  | IV | 57 | 110 | 11.0 | 92.9 | 146 |

We analysed each measure separately (head BPI + shoulder BPI + waist BPI + hip BPI) related to sex (boys and girls), sexual maturation (stages I, II, III and IV).

When analysing height perception (head BPI), it revealed distinct patterns related to sex. A one-way ANOVA demonstrated a significant difference in height perception between sexes, with females exhibiting greater accuracy in height perception compared to males [F(1,98) = 15.9, p = 0.0001, η² = 0.139]. Furthermore, sexual maturation stage did not significantly affect head size perception [F(3,96) = 0.267, p = 0.849, η² = 0.008], indicating that perceived height remained consistent across different maturation levels. Prior to analysis, assumptions of normality (W = 0.992, p = 0.787) and homogeneity of variances [F(2,97) = 1.49, p = 0.230] were met.

A ANOVA examines the effects of sex and sexual maturation and their interaction on shoulder width perception (shoulder BPI). An analysis of variance (ANOVA) was conducted to examine the effects of sex and sexual maturation on the standardized shoulder index. For the effects of sex as assumption checks revealed that variance homogeneity was met (Levene’s test: F(1,98) = 2.51, p = 0.116); however, the Shapiro-Wilk test indicated a significant deviation from normality (W = 0.953, p = 0.001) we transformed the raw data in zscore before conducting the ANOVA (Zscore_shoulder). The results indicated no significant effect of sex on Zscore_shoulder [F(1,98) = 0.074, p = 0.787, η²p = 0.001], suggesting that shoulder measurements did not differ between sexes. Similarly, assumption checks indicated a violation of variance homogeneity (Levene’s test: F(3,96) = 4.38, p = 0.006) and normality (Shapiro-Wilk: W = 0.966, p = 0.011) and ANOVA did not reveal a significant effect of sexual maturation on Zscore_shoulder [F(3,96) = 0.825, p = 0.483, η²p = 0.025], suggesting that shoulder measurements remained stable across different maturation stages.

An ANOVA was performed to evaluate the effects of sex and sexual maturation and their interaction on waist width perception (waist BPI). As assumption checks revealed a violation of homogeneity of variances for sexual maturation [Levene’s test: F(3,96) = 4.00, p = 0.010] and sex [Levenes’s test: F(1,98) = 5.32, p = 0.023], and the normality assumption was violated for sexual maturation [Shapiro-Wilk: W = 0.959, p = 0.003] and sex [Shapiro-Wilk: W = 0.952, p=0,001], we transformed the raw data of waist BPI in zscore and then conducted the ANOVA. The results showed no significant effect of sexual maturation [F(3,96) = 1.20, p = 0.315, η²p = 0.036], indicating that waist width body perception z-scores did not differ significantly across maturation stages. Similarly, there was no significant effect of sex and waist width body perception [F(1,98) = 2.42, p=0,123].

A ANOVA was conducted to assess the effect of sexual maturation and sex on hip width perception (hip BPI). Assumption tests revealed violations of homogeneity of variances for both sexual maturation [Levene’s test: F(3,96) = 4.98, p = 0.003] and sex [Levene’s test: F(1,98) = 5.17, p = 0.025], and the normality assumption was violated for sexual maturation [Shapiro-Wilk: W = 0.938, p < 0.001] and sex [Shapiro-Wilk: W = 0.919, p < 0,001], therefore we transformed the raw data in zscore..The results indicated a marginal effect of sexual maturation [F(3,96) = 2.65, p = 0.053, η²p = 0.076], suggesting a trend toward significance but not meeting the conventional threshold. Similarly, no significant difference was found between sexes [F(1,98) = 0.708, p = 0.402, η²p = 0.007]. Figure 2 shows the mean and confidence intervals of oBPI, head BPI, shoulder BPI, waist BPI, hip BPI related to sexual maturation (stages I, II, III and IV).

**Figure 1.**
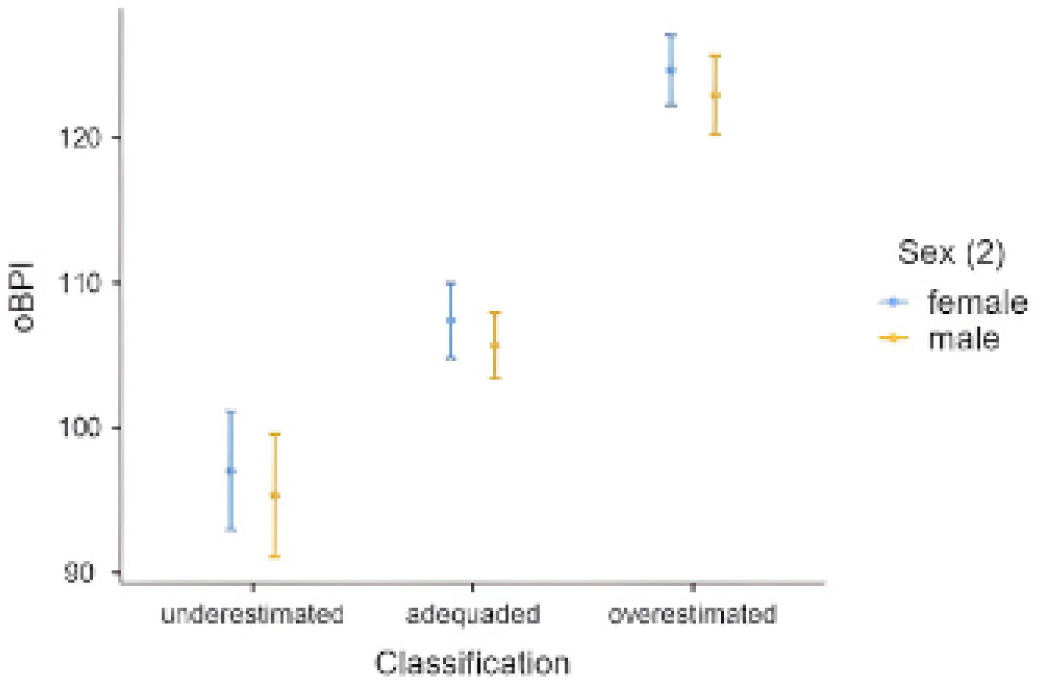
Presents the overall Body Perception Index (oBPI) in females and males.

**Figure 2.**
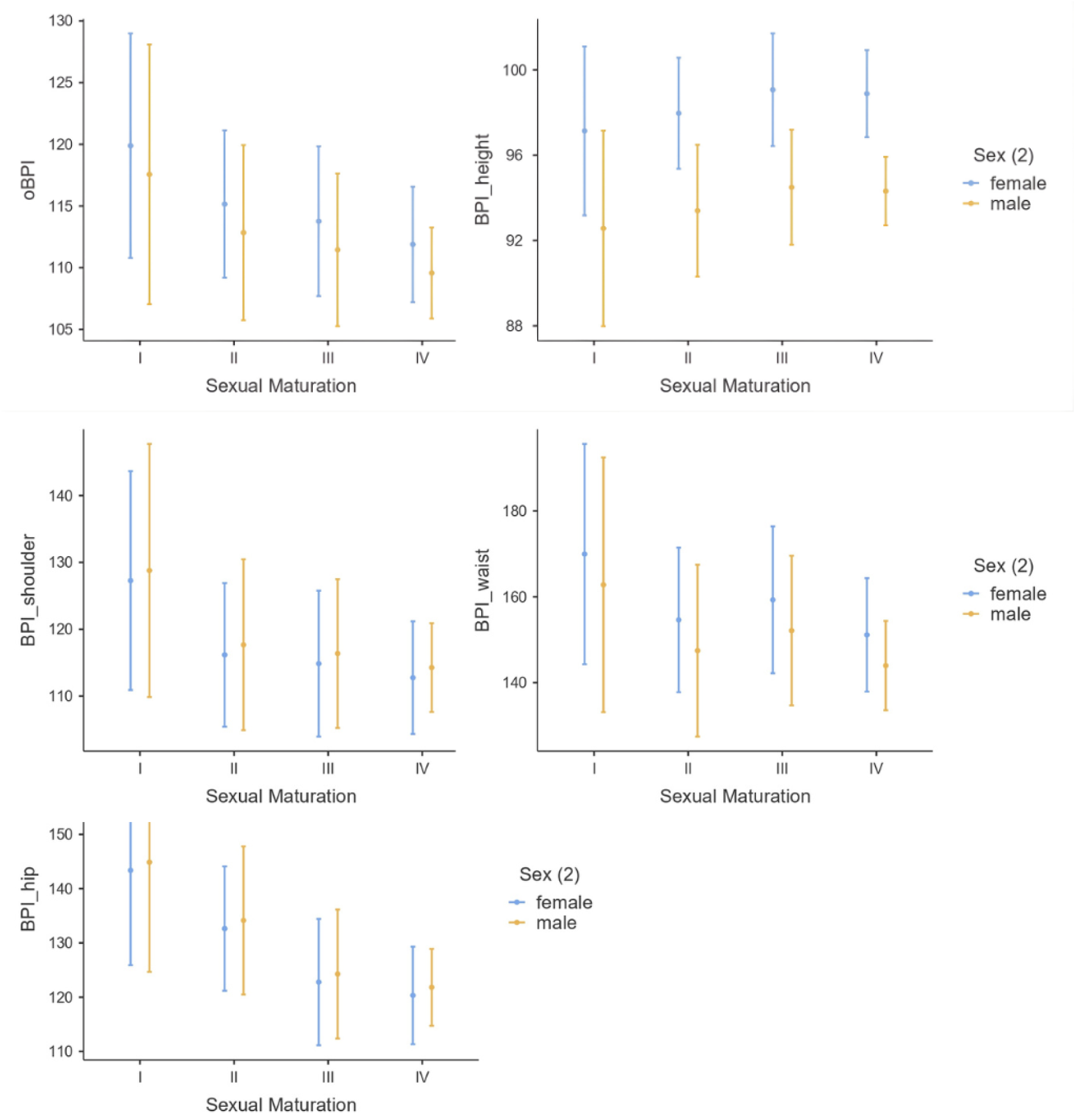
Mean and confidence intervals of oBPI, head BPI, shoulder BPI, waist BPI, hip BPI related to sexual maturation (stages I, II, III and IV) in males and females.

Descriptive statistics revealed sex differences in Previous Motor Activities. Males (mean = 18.8, SD = 32.9) exhibited significantly higher mean activity levels and greater variability compared to females (mean = 8.82, SD = 15.5). Analysis across sexual maturation stages showed a numerical trend of increasing mean motor activity from Stage I (mean = 4.71, SD = 5.96) to Stage IV (mean = 18.9, SD = 32.6), accompanied by increasing variability.

A one-way ANOVA on z-score transformed data, conducted to examine the effect of sexual maturation on Previous Motor Activities, yielded no statistically significant effect (F(3, 96) = 1.87, p = 0.139), despite the observed numerical increases across stages. Similarly, while an ANOVA investigating the effect of sex on z-scored Previous Motor Activities approached statistical significance (F(1, 98) = 3.76, p = 0.055), violations of homogeneity of variances and normality assumptions limit the interpretability of this trend, which suggested lower activity scores in females (mean difference = -0.382, p = 0.055).

Finally, a one-way ANOVA on z-score transformed data found no significant main effect of body size perception classification (underestimation, adequate, overestimation) on z-scored Previous Motor Activities (F(2, 97) = 0.218, p = 0.805).

In summary, while descriptive analyses revealed higher and more variable previous motor activity levels in males and a numerical increase across pubertal stages, subsequent inferential statistics on transformed data did not yield statistically significant effects for sexual maturation or body size perception. A trend towards lower motor activity scores in females approached significance but was limited by violations of statistical assumptions. These findings suggest that while initial observations point to potential associations, further research with larger samples and methods robust to non-normality is needed to definitively establish the relationships between sex, pubertal development, body size perception, and previous motor activities in this adolescent population.

Following the examination of Previous Motor Activities, we also investigated Global Motor Inactivity. Descriptive statistics for Global Motor Inactivity revealed a trend of increasing mean passive activity with advancing sexual maturation, ranging from a mean of 10.7 (SD = 4.96) in Stage I to 26.2 (SD = 15.6) in Stage IV, with considerable individual variability, especially in later stages. Regarding sex differences, males exhibited slightly higher mean passive activity (M = 25.4, SD = 14.8) compared to females (M = 21.5, SD = 14.2), although both groups showed similar variability.

An ANOVA on z-scored Global Motor Inactivity by body size perception classification (underestimation, adequate, overestimation) showed no statistically significant main effect (F(2, 97) = 2.420, p = 0.095, ηp2=0.047).,

The ANOVA examining the effect of sexual maturation on z-scored Global Motor Inactivity yielded a statistically significant main effect (F(3, 96) = 2.890, p = 0.040). Post hoc analysis indicated significantly lower inactivity z-scores in Stage IV compared to Stage I (p = 0.038), suggesting an inverse relationship between pubertal progression and passive activity. However, violations of homogeneity of variances and normality necessitate careful interpretation of this finding. We also investigated the effect of sex on z-scored Global Motor Inactivity with ANOVA showed no statistically significant main effect (F(1, 98) = 1.80, p = 0.182).

In summary, descriptive analyses indicated an increase in global motor inactivity with advancing sexual maturation and slightly higher levels in males. While inferential statistics revealed a significant effect of sexual maturation on inactivity z-scores, with higher maturation associated with lower inactivity, this finding should be interpreted cautiously due to violations of ANOVA assumptions. No significant effects of body size perception or sex on global motor inactivity z-scores were found, although a trend suggested higher inactivity in the underestimated group. The consistent violation of normality across several analyses highlights the need for careful consideration of data distribution in future investigations of these relationships within adolescent populations.

## 4. Discussion

This study explored the perception of body size across sexes and stages of sexual maturation using a projective test, revealing a significant overestimation of overall body size in girls that tends to diminish with pubertal progression, alongside a consistent distortion in waist width perception across the sample. Our analysis of body perception in adolescents revealed a significant tendency for girls to overestimate their overall body size, while boys demonstrated a more accurate perception. Notably, this overestimation in girls appears to diminish as they progress through sexual maturation, suggesting a dynamic interplay between body image and pubertal development. When examining specific body dimensions, girls exhibited greater accuracy in perceiving their height compared to boys. Notably, waist width emerged as the most distorted measurement across the entire sample, with no significant differences observed between sexes or across stages of sexual maturation for shoulder and waist width perception. Hip width perception showed a marginal trend towards being influenced by sexual maturation, as shown in Figure 2.

Our data shows that girls, as a group, overestimate their body size (48%) and most boys present an adequate perception of their body (58%), while only a small portion of boys underestimate (10%). The persistent overestimation in girls likely reflects the internalization of societal beauty standards that often emphasize thinness, leading to a discrepancy between their actual and perceived body size. Boys, on the other hand, may experience different pressures, such as those related to muscularity, which might result in a more balanced overall perception in our sample.

Our finding of body size overestimation in girls contrasts with studies in younger Brazilian children (Costa et al., 2015) and Korean adolescents (Chae, 2022), that reported different patterns and even underestimation in boys. In the Brazilian study, children from 7 - 10 years old showed that 34% underestimated and 42% overestimated their body size, with similar distribution between sexes. In the Korean study, most girls presented an overestimation while most boys presented an underestimation. These discrepancies likely stem from variations in age groups, cultural contexts, and the methodologies employed to assess body size perception. For instance, the emphasis on thinness in Western cultures during adolescence, as opposed to potentially different ideals in younger children or Korean adolescents, could contribute to these distinct patterns. Similarly, underestimation of body size in boys has been reported in Norwegian studies using the Children’s Body Image Scale (Steinsbekk et al., 2017) and in groups at high risk of eating problems (Sand et al., 2011), as well as in Israeli boys (Ben-Yaish et al., 2021). These findings, along with observations of underestimation in overweight children across multiple countries (Quick et al., 2014), highlight the complex interplay of age, culture, and individual characteristics in shaping body size perception. Interestingly, a Dutch study indicated that body size overestimation in girls becomes more pronounced during adolescence and persists into adulthood (Brug et al., 2006), suggesting that the sex differences observed in our adolescent sample may continue into later life. This reinforces the idea that these sex-related variations are not established in adulthood but rather take shape during the initial phases of sexual maturation and are likely to endure into later life.

The greater accuracy in height perception among girls is an interesting finding,, possibly because height is a more objective attribute less subject to social pressures compared to body width dimensions. The consistent distortion of waist width across all sexes and maturational stages powerfully underscores the pervasive influence of sociocultural ideals emphasizing a slim waist as a marker of attractiveness during adolescence (Dittmar et al., 2009). This finding highlights the strength of these societal pressures on body image, potentially overriding developmental changes.

While the overall effect of sexual maturation on body perception did not reach statistical significance, the trends observed in the Stage I female subgroup, with higher mean oBPI and variability, suggest that early puberty might be a period of greater instability in body image. This warrants further investigation in larger, more balanced samples to clarify the potential nuances of this relationship অনুযায়ী Coleman & Hendry, 1999).

The lack of significant effects of sex and sexual maturation on shoulder and waist width perception suggests that sociocultural ideals may exert a stronger influence on the perception of these specific body dimensions than the biological changes associated with puberty. The consistent distortion in waist perception, in particular, across all stages and both sexes, highlights the potency of societal pressures related to this body area. The marginal trend observed for the influence of sexual maturation on hip width perception hints at a possible subtle effect of pubertal body shape changes, particularly in females. Future research with larger samples is needed to determine the statistical significance and the nature of this relationship.

Our study’s novel approach, employing a projective test (IMP) to assess actual body size perception across different stages of sexual maturation provides valuable insights. Notable, we identified some individuals exhibiting significant overestimation (e.g., females in Stage I with oBPI of 162%; and in Stage IV with oBPI of 145%; and males in Stage III and IV, with oBPI of 130% and 132% respectively) or underestimation (e.g., a female in Stage IV with oBPI of 92%, a male in Stage III with oBPI of 93%), highlighting the variability in body perception within the adolescent population. Given the well-established link between body size overestimation and eating disorders (Brown et al., 2021; Gardner & Brown, 2014; Garner & Garfinkel, 1981) our findings underscore the potential utility of the IMP as a screening tool to identify adolescents who may be at higher risk and warrant further investigation for other risk factors (Barakat et al., 2023). The substantial overestimation found in young patients with eating disorders compared to controls (Schneider et al., 2009) further emphasizes the While body image distortion is a multidimensional construct encompassing perceptual, sensorimotor, and proprioceptive aspects (Gaudio et al., 2014, p. 22–14, 2014; Riva, 2012) encompass perceptual, sensorimotor, and proprioceptive dimensions. While some researchers argue that attitudinal aspects of body image are more crucial than perceptual ones in predicting treatment outcomes (Fernández-Aranda et al., 1999), our findings, along with those of Sand et al. (2011), suggest that perceptual distortions are indeed significant and may play a crucial role in the development of body image disturbances and potentially eating disorders. Therefore, addressing these perceptual biases early on could be key to preventative interventions aimed at promoting realistic body image and healthy attitudes towards their bodies.

We propose that the IMP, as an accessible and cost-effective projective test that taps into somatosensory processing and body awareness (Mandrigin, 2021), holds promise as a screening tool for identifying children and adolescents who may be more susceptible to developing eating disorders. Its ease of use allows for integration with other attitudinal measures of body image (Thurm et al., 2011). The finding that body size perception patterns observed in adolescence appear to align with those in young people and adults suggests that early intervention strategies targeting these distortions could prevent their persistence.

Although our findings did not reveal a direct influence of prior motor activities on body size perception across pubertal stages, the literature suggests a positive association between physical activity and body image satisfaction (Gualdi-Russo et al., 2022; Toselli et al., 2022; Zaccagni & Gualdi-Russo, 2023). Therefore, promoting healthy engagement in sports and physical activity, while being mindful of potential risks (Burgon et al., 2023), remains a valuable intervention strategy for overall well-being and potentially for mitigating body image dissatisfaction that can accompany perceptual distortions.

We believe that the IMP, as a projective test, could be used as a screening tool to investigate children and adolescents who might be more likely to develop eating disorders. It is an easy and cheap which involves localizing a touched point on the body surface which elicit a somatosensory process to identify the stimulus in the somatotopic map and the integration with memory and motor areas to project your perceived height, shoulder (acromion), waist (last rib) and hips (trochanter) position. We believe that the IMP requires a cross-modal frame from sensory reference and motor output (Mandrigin, 2021), providing a better bodily awareness representation assessment. The IMP has been suggested as an easy tool to be used with other attitudinal aspects of body image evaluation in eating disorders (Thurm et al., 2011).

Since our study showed that the alterations in the perception of body size manifested in adolescence/infancy correspond to the pattern of body perception in young people and adults, we can think of strategies that interfere with this distortion in its early stages as a way of preventing these alterations from continuing.

One of these strategies could be to encourage physical exercise. In our findings, the previous motor activities and global motor inactivity seems to not influence the body size perception in different stages of sexual maturation, but related to body image dissatisfaction, it was found that Italian adolescents showed a higher level of body image dissatisfaction in non-sports-playing females, while sports-playing males showed the lowest dissatisfaction (Toselli et al., 2022). It seems like as the physical activity increases in adolescence, the body image dissatisfaction decreases (Gualdi-Russo et al., 2022). In adulthood, athletes are generally believed to be more satisfied when compared with the non-sports population (Zaccagni & Gualdi-Russo, 2023). Promoting healthy engagement in sports and physical activity, while being mindful of the potential risks of excessive exercise (Burgon et al., 2023), could be a valuable strategy.

Furthermore, interventions addressing the influence of social media and societal pressures on body image are crucial. Reducing exposure to idealized images (Friederich et al., 2007; Huang et al., 2021; Kleemans et al., 2018; Thai et al., 2024; Vincente-Benito & Ramírez-Durán, 2023), promoting body positivity (Rajesh & Draper, 2022). and developing critical thinking skills regarding media content (Cortez et al., 2016; Mahon & Seekis, 2022) are essential.The development of a positive body image and a healthy self-esteem can be stimulated by parents, teachers and clinicians (Hartman-Munick et al., 2020; Tort-Nasarre et al., 2023).

It is important to consider the limitations of this study while interpreting the findings. First the low participation rate limits the generalization of this sample. We did not evaluate the data related to eating disorders and other potential psychological factors that could be influencing the perception of body size, which future studies should address. The cross-sectional design limits our ability to establish causal relationships between sexual maturation and body perception changes. The reliance on self-report measures for body perception may also introduce subjective biases. Furthermore, the sample size for some sexual maturation stages, particularly Stage I, was relatively small and exclusively female, which may have limited the statistical power to detect significant effects and the generalizability of findings related to sexual maturation. Future research should employ longitudinal designs with larger and more diverse samples to investigate the developmental course of body perception across puberty.

## 5. Conclusion

This study delivers compelling insights into the intricate development of body size perception during adolescence, demonstrating a significant tendency towards overestimation, particularly pronounced in girls and during the initial stages of sexual maturation.

This study provides valuable insights into the development of body size perception during adolescence. The observed trend towards overestimation, particularly in girls and in the early stages of sexual maturation, highlights the need for early identification and intervention. The IMP may offer a useful screening tool for identifying at-risk individuals. Future research should investigate the longitudinal trajectory of body size perception and its relationship with the development of eating disorders. Interventions targeting both individual and societal factors, including promoting healthy physical activity, critical media literacy, and positive body image, are essential for preventing body image disturbances and promoting well-being in adolescents. Further research exploring the neuropsychological underpinnings of the IMP and its specific relationship to various dimensions of body image is warranted.

In conclusion, our study confirms the well-established pattern of overall body size overestimation in adolescent girls and highlights the unique accuracy in their height perception. The persistent and consistent distortion of waist width perception across sexes and maturational stages underscores the powerful influence of sociocultural ideals on body image during this developmental period. Although the overall influence of sexual maturation on body perception in our sample did not reach statistical significance, the observed trends, coupled with the marginal effect on hip perception, strongly suggest a more nuanced and complex interplay deserving of rigorous investigation through longitudinal studies employing larger, more balanced samples. Ultimately, these findings unequivocally highlight the critical and immediate need for targeted interventions that address both gender-specific body image concerns and the pervasive societal pressures that contribute to detrimental body perception distortions in adolescents, thereby safeguarding their present and future well-being.

## Data Availability

All data produced in the present study are available upon reasonable request to the authors

